# An analysis of patients’ perspectives on qualitative olfactory dysfunction using social media

**DOI:** 10.1101/2020.12.30.20249029

**Authors:** Jane K. Parker, Christine E. Kelly, Barry Smith, Claire Hopkins, Simon B. Gane

## Abstract

**Background:** The impact of qualitative olfactory disorders is underestimated. Parosmia is the triggered perception of distorted odours whereas phantosmia is the perception of odours in the absence of a trigger. Both can arise from post-infectious anosmia and have increased substantially since the outbreak of COVID-19.

**Methodology/Principal:** Thematic analysis of a social media support group for parosmia and phantosmia was used to articulate the perspectives and concerns of those affected by these debilitating olfactory disorders.

**Results:** A novel symptom (olfactory perseveration) was identified where a triggered, identifiable, and usually unpleasant olfactory percept persisted in the absence of an ongoing stimulus. Fluctuations in intensity and duration of perseveration, parosmia and phantosmia were observed. Coffee, meat, onion, and toothpaste were identified as common triggers of these disorders, but people struggled to describe the distortions, using words associated with disgust and revulsion. Common strategies to avoid triggers may result in a diet lacking in both nutrition and reward. The emotional aspect of living with qualitative olfactory dysfunction was evident and highlighted the detrimental impact on mental health.

**Conclusions:** The data acquired can inform rehabilitation strategies and drive our ongoing research into understanding the molecular triggers associated with parosmia, and research into patient benefit.

## INTRODUCTION

Until recently, olfactory dysfunction was a little-recognised and underestimated disorder, distressing to those affected and with few effective treatments available. Prior to the COVID-19 pandemic, olfactory dysfunction was believed to affect about 5% of the general population^1^ rising to 20% of those aged over 60. However a more recent meta-analysis gave an estimated prevalence of 22%^2^ when objective measures were employed^3-4^. The aetiologies most commonly reported are sinonasal disease, upper respiratory tract infection and traumatic brain injury^5^.

Since the outbreak of the COVID-19 pandemic, cases of olfactory dysfunction have increased. The most recent estimate for COVID-19-related loss of smell and taste is 65%^6-7^. Given that there have been 80 million cases of COVID-19 globally (to the end of 2020)^8^, several million people will have been affected by smell loss. Although many will recover within weeks^9^, it is estimated that about 10% will have long term olfactory problems, many of whom will subsequently develop a qualitative olfactory dysfunction^10^. The term qualitative olfactory dysfunction covers both parosmia (qualitative distortion in the presence of an odour), and phantosmia (odour experience in the absence of an odour).

Parosmia is the triggered (requiring an external stimulus), subjective perception of a qualitatively altered odour identity with negative hedonic component (almost universally unpleasant) which usually subsides within seconds of the stimulus. It often develops in the early stages of recovery from smell loss, particularly after post-infectious and post-traumatic anosmia, with onset weeks after the initial insult. Those severely affected by parosmia find many familiar food aromas intolerable and start to reject food, leading to weight loss, anxiety and in severe cases to clinical depression^11-12^. Parosmia is reported in 34% of all patients presenting with olfactory disorders (n=392)^13^ and the most common aetiology is upper respiratory infection, with 56% of those with anosmia progressing to parosmia.

Phantosmia often occurs alongside parosmia^14^ and is similarly a perception of an unpleasant subjective odour but is not triggered by obvious external odorants. Patients experience many of the same objectionable odours that are perceived by parosmics^5^ but these sensations can persist for days.

Awareness of anosmia has undoubtedly increased since the start of the COVID-19 pandemic, after it became recognised worldwide as one of the key symptoms of COVID-19^15^. However, relatively few publications are dedicated to understanding both the pathophysiology of the disease and its impact on the patient. The charity AbScent (Registration No. 1183468 in England and Wales) has provided support for those with such disorders, launching several support groups on Facebook since 2018: a COVID-19 Smell and Taste Loss group was started in March 2020 and, once it became apparent that post-COVID-19 anosmics were also developing parosmia, a group dedicated to those experiencing parosmia and phantosmia, was started in June 2020. This group accumulated over 4700 followers by December 2020 and is an important source of information for researchers, providing valuable insight into the nature and progression of the disease.

The fact that people are turning to social media for support is evidence that patients are in need of more information to help them understand both the disease and the efficacy of various treatments, as well as find social interaction and moral support in coping with an often debilitating condition. In this paper we use qualitative data from the AbScent Parosmia and Phantosmia support group to understand the underlying themes of parosmia and phantosmia, the concerns of those afflicted and the nature of the foods that trigger the distortions. Understanding these will support further research into the mechanisms and therapeutic options for this condition.

## MATERIALS AND METHODS

This study has been given University of Reading School of Chemistry, Food and Pharmacy Research Ethics Committee approval (study number 39.2020) and permission has been given for use of all personal quotes.

The use of social media is something that appears rarely in the published literature, yet Alanin et al^16^ argue strongly that this approach, driven by the patients’ own perspectives, provides invaluable, unsolicited and spontaneous data that would not otherwise be retrieved from more structured surveys and questionnaires. It paints a colourful picture of the patient journey, avoiding bias arising from the structure of the survey and the generation of artefacts where patients are overeager to please. Other biases may be introduced with this method, for example, the sample is not randomised and no conclusions about incidence or prevalence can be made due to the self-selection of this group. While much of the research into smell and taste changes during the pandemic has focused on patients who have had confirmed cases of COVID-19, either through positive tests or clinical diagnosis, the information presented here is about recent self-reported changes which cannot be confirmed to be related to COVID-19 infection.

The findings reported here are taken from the AbScent Parosmia and Phantosmia Support closed group on Facebook between 12^th^ June and 14th December 2020. Conversation within this group is lively, and responses to polls and questions can generate over 300 comments in 24 h.

### Thematic Analysis

The Facebook group moderator (CK) monitored the discussion daily, noting down recurring themes, and a second researcher (JP) independently reviewed the posts to identify themes. The lists were combined and after discussion, 7 major themes were identified by consensus and unattributed quotes were selected to support the thematic groups. Subthemes emerged, and 1-2 quotes for each sub-theme were selected for presentation in Table 1.

**Table 1.**
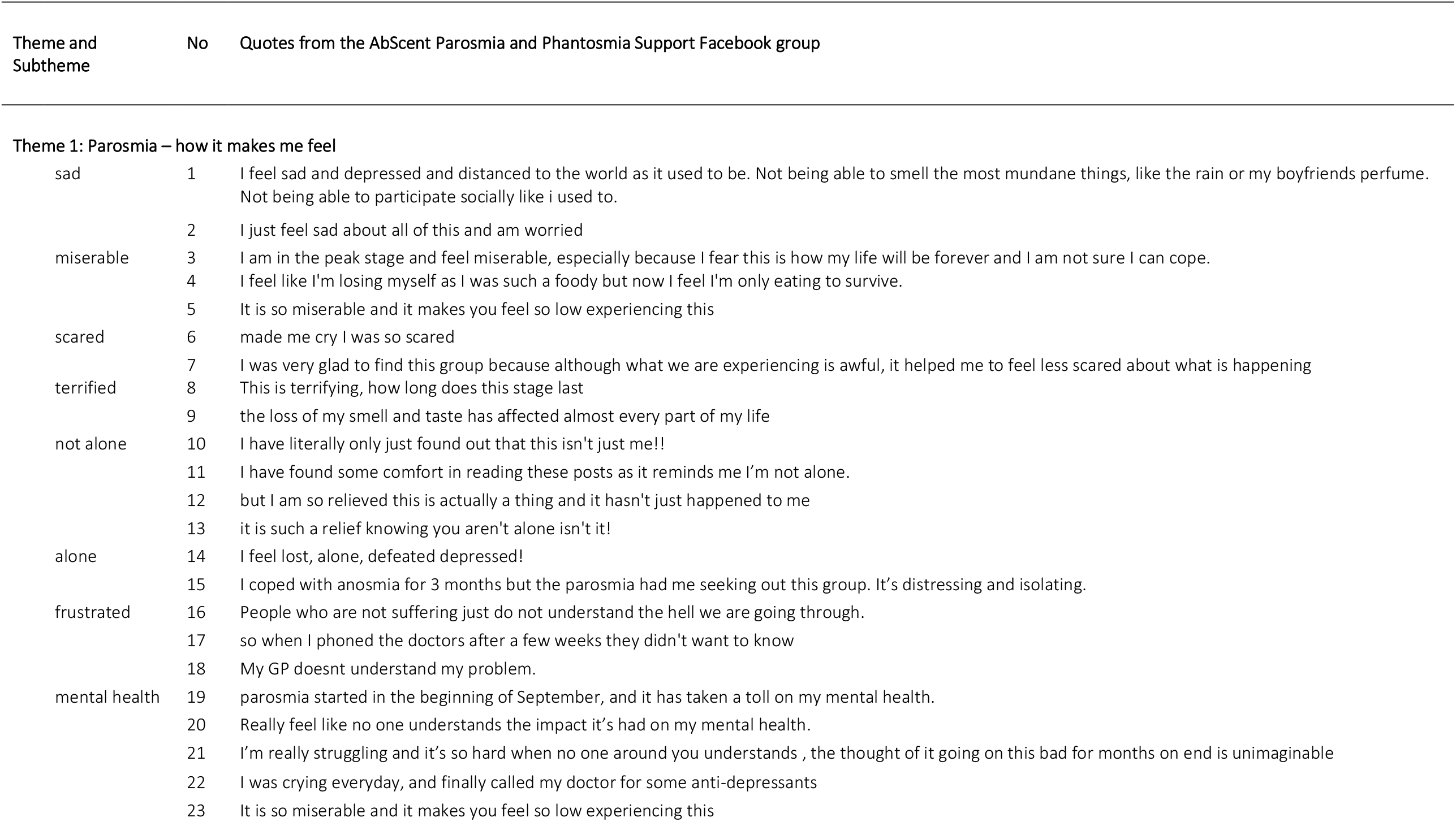
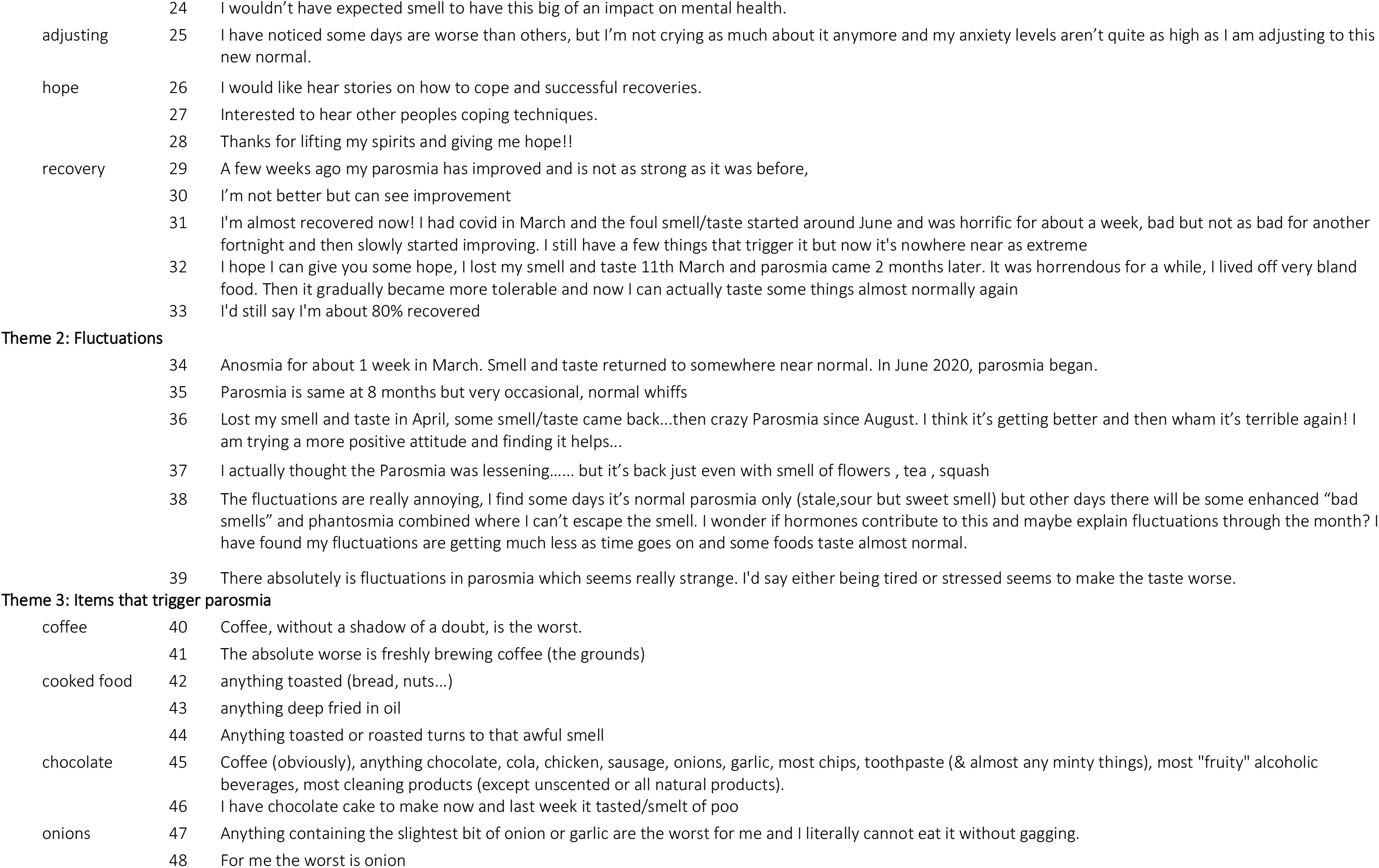
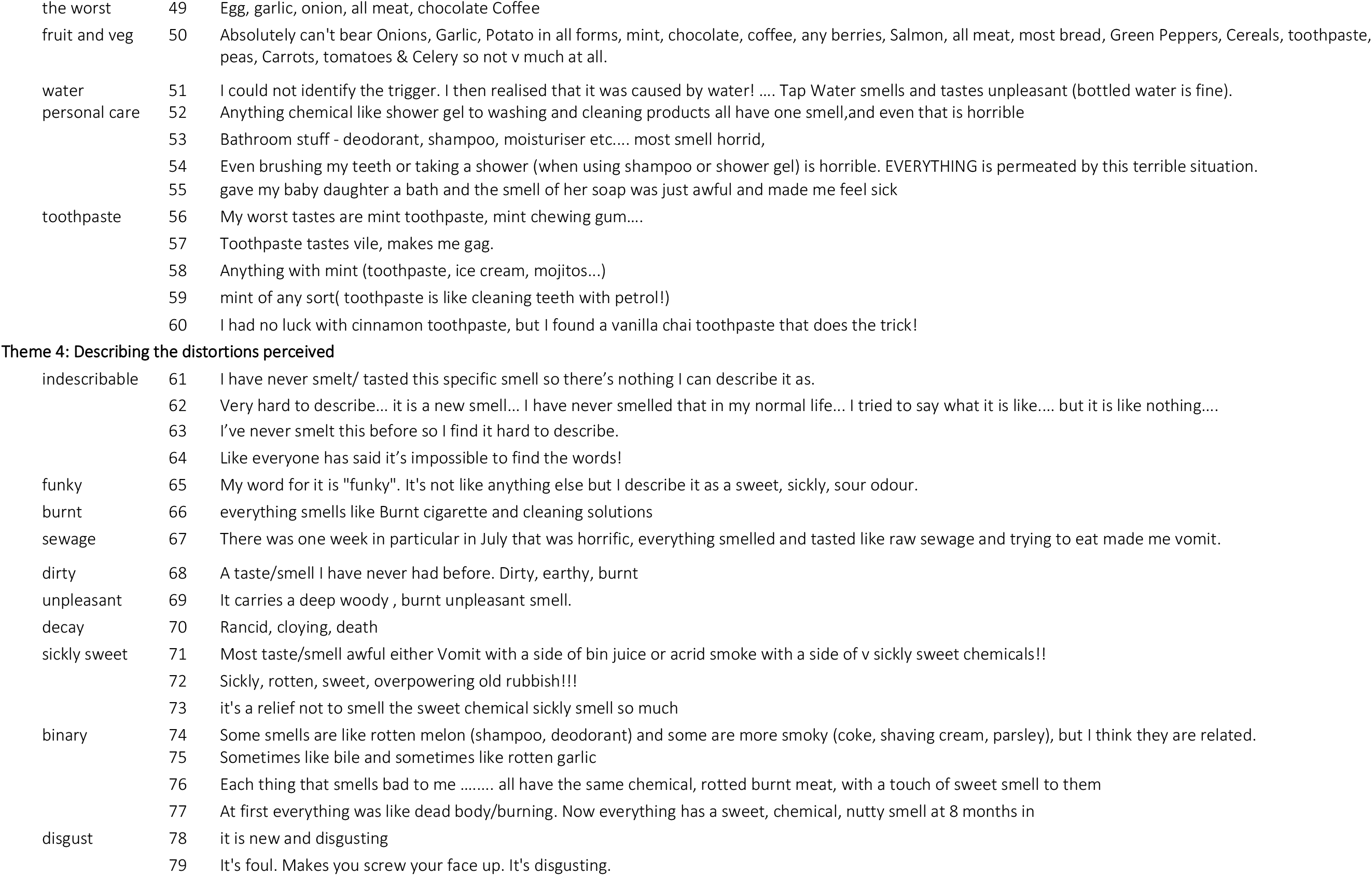
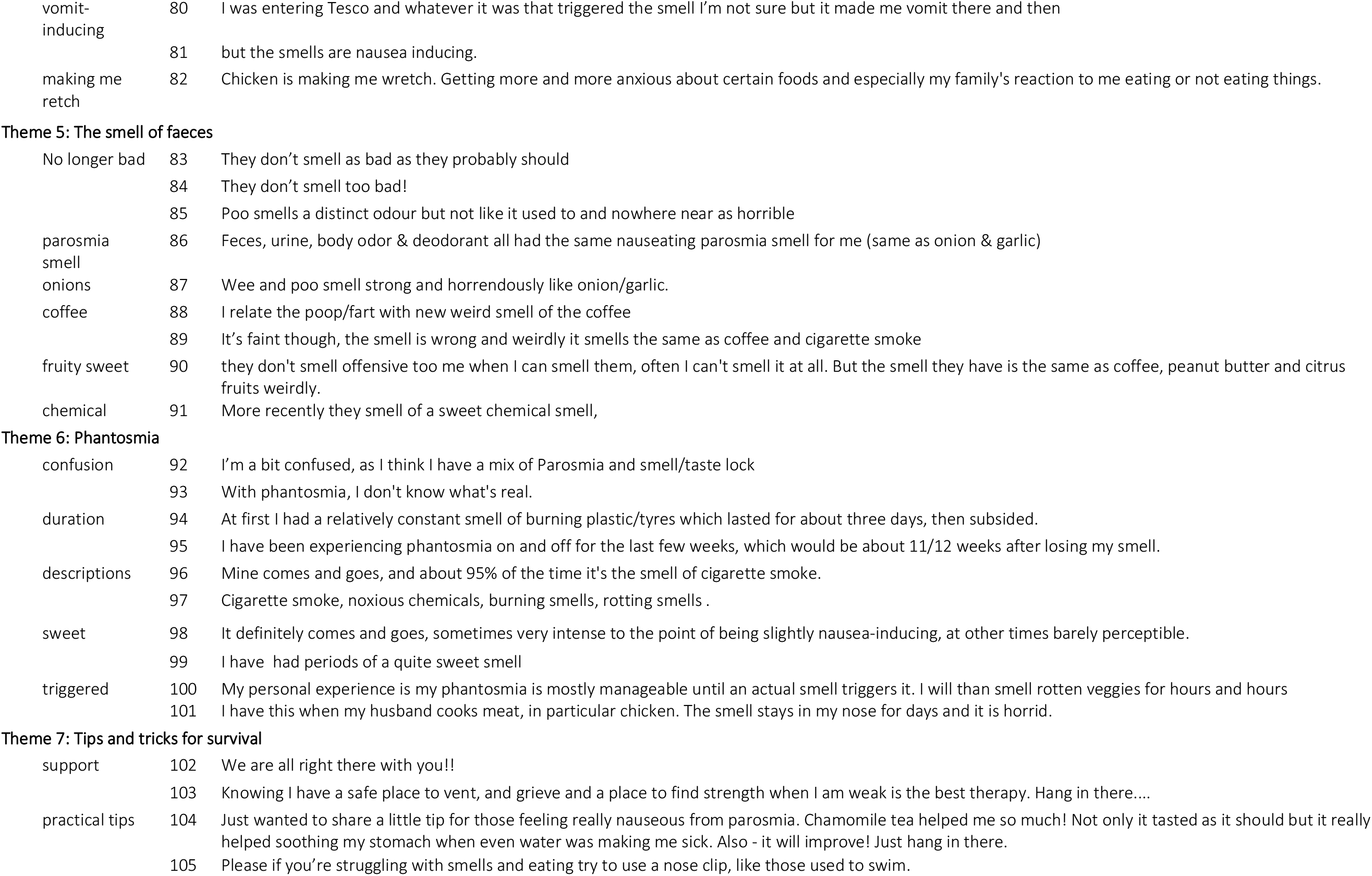
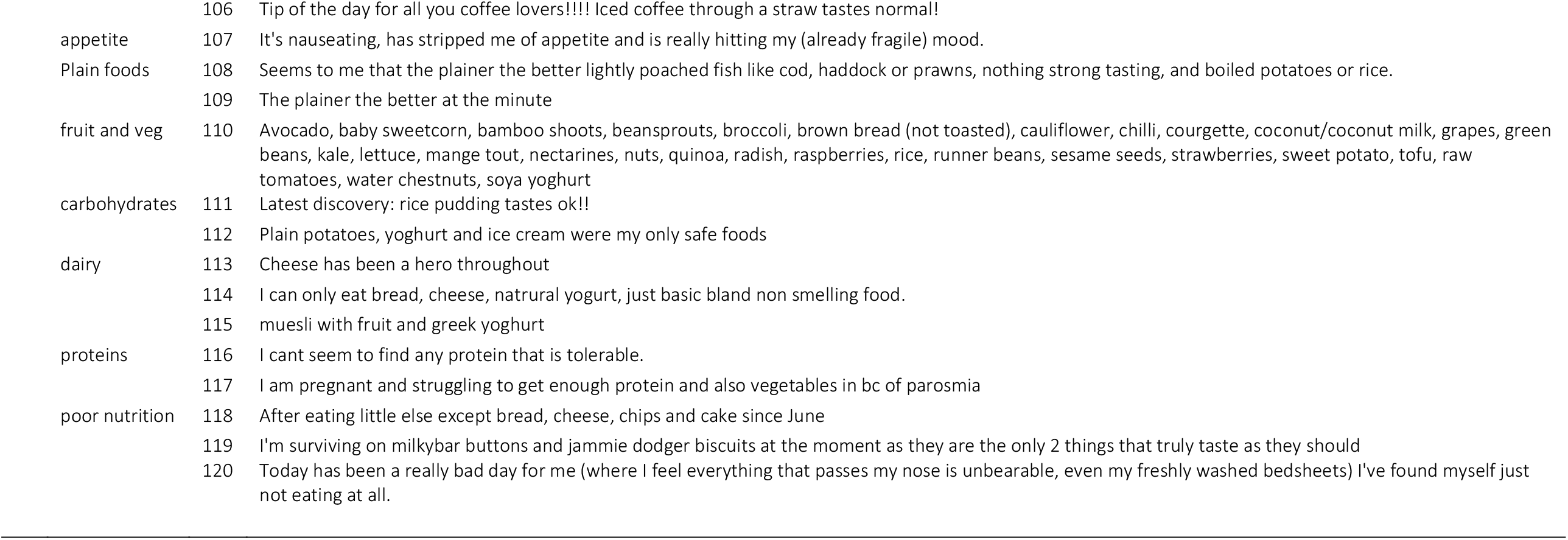
Extracts from the AbScent Parosmia and Phantosmia Support Facebook group

### Content analysis

To identify the foods most often associated with parosmia, the thread of one conversation was analysed manually. The conversation was prompted by the moderator who posed the question: “Can you all add here your worst foods for parosmia?” (137 comments). Data were collected during the first three months of the study using a simple frequency table to record each food as it was mentioned (Table 2). Some items required grouping, for example, the generic term “meat” was used, forcing us to combine this with chicken, beef, lamb, and pork. Sausages, being a mixture of meat, onions, garlic and spices were not included and bacon was considered different based on the curing process and the underlying chemistry. The terms lime, lemon and orange were grouped with citrus, and all personal care products gathered under one term. In general, complex food items containing a number of potential triggers (curry, fish fingers, sausages) were excluded from the count. Mint and toothpaste were combined. In terms of text analysis, this is a very small data set, and manual coding was deemed appropriate, although the repeatability and tolerance associated with this method should be borne in mind.

**Table 2.** Count of Trigger Foods Reported taken from 137 comments retrieved from the AbScent Parosmia and Phantosmia Support Facebook group between 8^th^ August and 30^th^ September 2020

| **Cooked foods** |  | **Fruits, vegetables,** |  | **Alliums/Brassica** |  | **Drinks** |  |
| --- | --- | --- | --- | --- | --- | --- | --- |
| **Maillard** |  | **herbs and spices** |  |  |  |  |  |
| meat (chicken, beef, lamb) | 45 | banana | 14 | onion | 47 | wine red and white | 14 |
| coffee | 42 | citrus | 13 | garlic | 27 | cola | 11 |
| eggs | 28 | peppers | 6 | mustard | 3 | water | 1 |
| chocolate | 14 | cucumber | 6 | rocket | 2 |  |  |
| fried foods | 11 | coriander/cilantro | 6 | broccoli | 1 | **Non-food items** |  |
| peanuts/peanut butter | 9 | berries | 5 |  |  | toothpaste/mint | 43 |
| bread | 7 | strawberries | 4 | **Dairy** |  | personal care products | 30 |
| bacon | 6 | celery | 3 | butter | 3 | body odours | 5 |
| toast | 5 | jalapenos | 2 | cheese | 3 | cigarette smoke | 4 |
| tomato products | 4 | pineapple | 2 | yogurt | 2 | bleach | 4 |
| humous | 3 | pesto | 2 | milk | 1 | petfood | 3 |
| oats/porridge | 3 | watermelon | 2 |  |  | petrol | 2 |
| soy sauce | 2 | spices | 2 | **Flavoured products** |  |  |  |
| marmite | 2 | parsley | 1 | sweets | 3 |  |  |
| popcorn | 1 | peaches | 1 | crisp flavours | 3 |  |  |
|  |  | spinach | 1 | vanilla | 3 |  |  |
|  |  | basil | 1 |  |  |  |  |
|  |  | ginger | 1 |  |  |  |  |
|  |  | apple | 1 |  |  |  |  |

## RESULTS

### Theme 1. “How it makes me feel”

There is an extensive and concerning theme demonstrating the detrimental effect of parosmia and phantosmia on emotional well-being. Members not only report being sad and miserable (1-5), but some are quite scared and terrified by the prospect of having parosmia (6-9) and find reassurance when they find others in a similar position (10-13). Patients feel alone and isolated (14-15) and are relieved to find the support of the online community. Some are frustrated with lack of understanding from others (16) and responses from their doctors who seem uninterested (17-18) and lack treatment options. Many report a decline in their mental health (19-23) and one member was surprised that loss of smell could have such an impact on mental health (24). Lack of the ability to smell body odour is a worry as is the inability, from a safety perspective, to smell smoke or gas. Some adjust to the ‘new normal’ (25) and many are looking for hope (26-28). Although it is early days, some post about their recovery (29-33) which is usually only partial. Few mention complete recovery, but by this stage they may have left the group without commenting further.

### Theme 2. Fluctuations

One emerging theme is the fluctuating nature of olfactory dysfunction. Often there is partial (hyposmia) or complete (normosmia) return of a normal sense of smell before the onset of parosmia (34). Duration varies as some have occasional “whiffs” of normality occurring during long stretches of parosmia (35) but, for others, the changes in intensity and the magnitude of the fluctuations is quite extreme (36-38) and persistent (38) and the severity of parosmia and phantosmia (92-100) can vary daily. Both hormones (38) and tiredness or stress (39) were associated with worse fluctuations.

### Theme 3. Items that trigger parosmia

Much of the online discussion concerned key triggers, as patients started to recognise and record their own experiences and explore whether they shared the same triggers with others. Some list up to 15-20 different triggers which includes food, drinks, non-food items (bleach and cigarettes) and personal care items (shower gel, shampoo and hand sanitisers). Many mention coffee to be one of the worst (40-41). Fried, toasted or roasted foods are common triggers (42-44), as is chocolate (45-46) and onions and garlic (47-49) but these are not universal. Long lists of other foods are provided which include carbohydrates, fruit, vegetables, herbs, spices, (50) and even water (51), leaving it almost impossible to create a list of foods that are never triggers.

Personal care items are frequently mentioned (42-55), many attributing their parosmia to different brands, ingredients and explore alternative products. Toothpaste/mint are frequent offenders (56-59) and there are recommendations to switch to alternative flavoured toothpastes (60).

### Theme 4. Characteristics of the trigger foods

One defining feature of parosmia is the distortion of (familiar) smells which members struggle to describe as they cannot relate the smell to a previous experience (61-65). Although many descriptions have been used to describe “that parosmia smell”, they are often prefixed with “it’s like” in an attempt to describe the associated disgust, rather than the smell identity. A group of words frequently used seems to be based on a burnt, chemical or dirty connotation (burnt cigarettes, burnt rubber, sewage, earthy, dirty, rancid, death and decay, or unpleasant (66-70)), but there is evidence of at least one other type of parosmia smell with many reporting their triggers as sickly sweet and rotten (71-73). These two different concepts are often attributed to different foods (74-75) but have been used together to describe one parosmia smell (76) or in a progression where the burnt character appeared first but changed into the more sweet smell after several months (77).

The feeling of disgust and revulsion associated with the distortions (78-79) is clear and can induce vomiting in a handful of cases (80-82). Disgust is not always mentioned explicitly, but many imply their disgust by their choice of words (Theme 2) which are associated with disgust (garbage, sewage, decay, poo).

### Theme 5. The smell of faeces

This is a recurrent theme in this Facebook group. If perceived at all, the smell of faeces is usually more pleasant than expected (83-85) and often takes on the same character as food such as onion and garlic, whether this is distorted (86) or normal (87) onion and garlic. The smell of faeces is often described as distorted coffee (88-89) or as a sweet smell (90-91). The striking corollary of this is that the hedonic value of these odours is reversed: odours that typically elicit disgust are less objectionable than before, but odours which usually have a positive hedonic value are perceived as disgusting.

### Theme 6. Phantosmia

Many confuse parosmia and phantosmia (92), not knowing whether what they perceive is real (93). In general, phantosmia is discussed less frequently, even though it causes as much anxiety, as the perceived odour can last for days (94), weeks (95) or even months. Similar to parosmia, the descriptions of the smell are cigarette, chemical, burnt and rotting (96-97) or sweet and sickly (99), and phantosmia is subject to fluctuations (96, 98). An unusual and novel finding was that in some cases, members described it is a triggered reaction (100-101) and this was echoed strongly in the AbScent COVID-19 Small and Taste loss group where there was a thread dedicated to phantosmia.

### Theme 7. Tips and tricks for survival

Evidently the members of the group provide significant support for each other (102-103), creating a positive environment with few negative comments. Some post practical tips to mitigate the impact (104-106), and many are keen to pass on their own experience and provide lists of “safe” foods that cause minimal distortions. In general, it is no wonder that appetite is lost (107) as bland foods are frequently recommended (the plainer the better, 108-109)) and many turn to fresh fruit and vegetables (110), plain carbohydrates (111-112) and dairy products (112-115) as “safe” options. Cooked and roasted foods which tend to be a major source of protein (meat, nuts) are not considered “safe” and it is concerning that some struggle to find a palatable source of protein (116-117). Whereas some have found acceptable alternatives (quorn, turkey mince, and protein shakes) others report diets lacking in anything nutritious (118-119) or abstain from food completely (120), putting themselves at risk of malnutrition.

### Content analysis

More than 75 different items were mentioned as triggers in one conversation prompted by the moderator. By using our selection criteria (avoiding multi-ingredient foods, focusing on simple ingredients, and combining where necessary) a list of 50 items which trigger parosmia, and the frequency with which they were cited, is shown in Table 2. The four major food triggers, each cited >40 times, were coffee, meat, onions and toothpaste/mint, followed by garlic and eggs cited >20 times. Personal care products such as shower gel, deodorant, soap, shampoo, and hand sanitiser were often triggers, and these were gathered into just one category and mentioned >30 times. These results are displayed in a word cloud (Figure 1).

**Figure 1.**
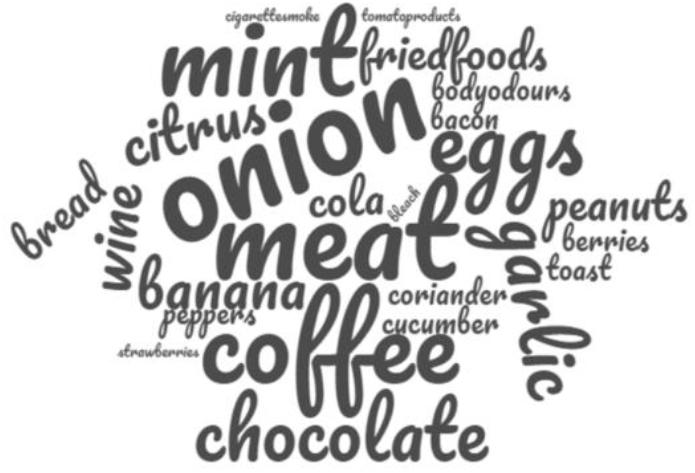
Wordcloud of common triggers where the size of the word is proportional to the frequency of citation.

## DISCUSSION

### Emotional Manifestations

The emotional and mental health impacts of smell disorders was a major topic of discussion in the group and is already well-documented in the literature^11-12, 17,18^. Figure 2 summarises the psychosocial manifestations, incorporating observations on food issues from Burges-Watson et al^18^ showing the interdependence of food issues and emotional effects. Smell loss or change is acutely felt by those suddenly confronted with the way their experience of the world is altered in terms of a lack of pleasure in eating and the absence of reassuring smells of familiar people and places. Many feel socially isolated and go on to suffer long lasting depression.

**Figure 2.**
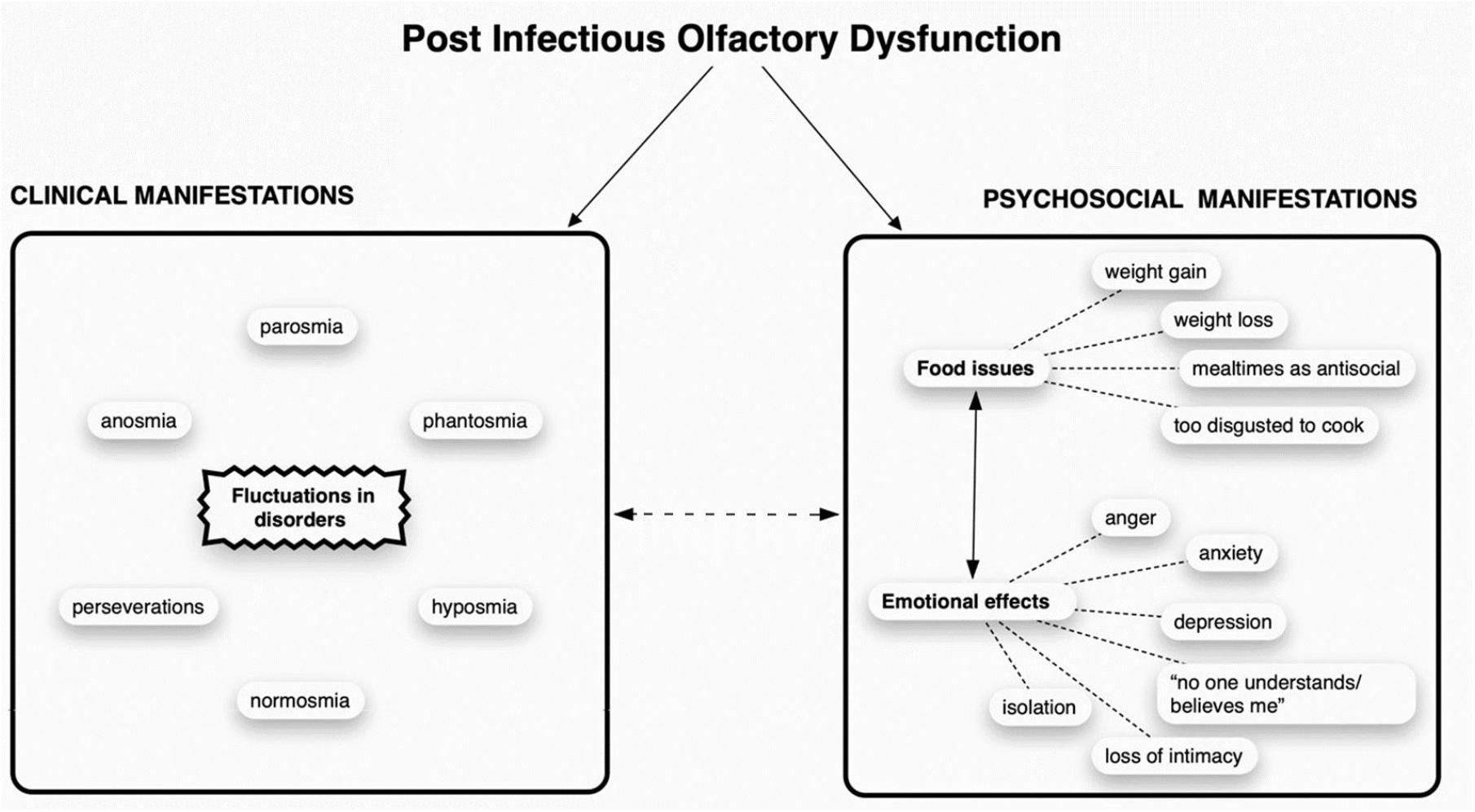
Clinical and Psychosocial Manifestations of Olfactory dysfunction.

### Clinical manifestations

Accounts of clinical manifestations are summarised in Figure 2 where there is an undefined yet complex and fluctuating relationship between different olfactory disorders (Theme 2). Contrary to what we have learnt anecdotally from non-COVID-19 incidences of parosmia, posts in this recent group indicate that parosmia can be preceded by the almost complete return of a normal sense of smell. This is hard to rationalise in terms of current olfactory neurobiology. During further progression of the disease, experiences of the symptoms of olfactory disorders vary in intensity and duration, (prolonged periods or short whiffs) giving episodes of different qualitative olfactory disorders and intermittent returns to anosmia or even what is perceived as a normal olfactory function. These “fluctuations” have no obvious basis, or sequence, can occur in series or in parallel, and vary from case to case. Given the close association between olfactory function and mental well-being^11^, it is conceivable that fluctuations may be influenced by emotional status.

What was surprising in Theme 6 was the elucidation of what we believe to be a novel symptom: ‘smell lock’ or olfactory perseveration. A search of the literature finds only one mention of this phrase before in a paper on olfactory hallucinations in Alzheimer’s disease^19^, where it was not further described. We propose a definition of this symptom as a triggered, identifiable, usually unpleasant olfactory percept that persists in the absence of an ongoing stimulus. We suspect that it is often confused with phantosmia and is relatively common in post-infectious smell loss syndrome. It is distinct from phantosmia in that it is triggered by a recognisable stimulus and remains identifiable as the smell of that trigger while persisting long after the original stimulus is removed from the surrounding environment. Describing novel symptoms such as this both increases our understanding of patient experience and guides further research efforts.

### Common triggers

The wide range of trigger food and personal care products identified in Theme 3 is consistent with a survey of 725 patients carried out by Keller et. al^5^ in 2013. This study also reported coffee as an “efficient” trigger and it was often mentioned together with chocolate. Meat, coffee, and cocoa represent some of the most complex aroma profiles, as the combination of sugars, amino acids and fats often exposed to high temperatures (coffee roasting, roasting of cocoa nibs, roast meat) produces a rich mix of both Maillard reaction (flavour forming reaction between sugars and amino acids) and lipid degradation products. Since these and other cooked foods listed in Table 2 (fried foods, peanut butter, bacon and toast) are frequent triggers which are rarely reported as “safe foods”, it is reasonable to suggest that they may have aroma compounds in common which trigger parosmia. Onion and garlic are mentioned far less frequently in the Keller et. al. study but are major triggers in this study. Both contain a variety of potent sulfur-compounds which we propose may be trigger molecules. However, brassica species (cauliflower, broccoli, kale) which also contain potent sulfur compounds are rarely mentioned, and indeed these three items appear in a list of “safe” foods (but not without exception). Many unheated foods are also triggers (bell pepper, citrus, apple, cucumber and banana) so parosmia is not simply associated with cooked foods. Mint and toothpaste, not mentioned by Keller et al., also seem to be powerful triggers, although it is not yet clear whether this might also be due to stimulation of the trigeminal nerve. Personal care products are objectionable to many, but these are very brand dependent and subject to the ingredients and particularly the choice of essential oils and flavourings used by different manufacturers.

In Theme 5 we find that the smell of faeces takes on the character of other distorted foods, suggesting that the strongly objectionable odours typically associated with faeces are not being perceived. There may be trigger compounds present in the faeces which are usually masked by the typical amino acid degradation products characteristic of faecal odour. There is a curious switching in hedonic valence between what are usually objectionable odours and those that are usually highly desirable: fair is foul, and foul is fair in parosmia, it seems.

### Coping Strategies

Advice on food is crucial both from a nutritional, hedonic and emotional perspective Theme 7). Avoidance of the top triggers (coffee, meat, eggs, onion, garlic and toothpaste) and most roasted or baked foods makes sense, but this may lead to nutritional deficiencies, especially in those who rely on meat and eggs as their main source of protein. Furthermore, avoidance of triggers may hinder the adaptation process which happens, albeit very slowly. This is one area where further research would be beneficial. It is far better for those afflicted to develop coping strategies for unpalatable foods. This could involve minimising thermal load, boiling or steaming rather than roasting or frying, and minimising flavour release by consumption at room temperature or chilled. Coping strategies may also include avoidance of grocery stores where the smell can be overwhelming and ordering food online for delivery. What is clear is that every person is different, and people need to experiment to find a varied diet with the right balance of nutrition and reward.

## Conclusion

Qualitative methods are sometimes perceived as being ‘less rigorous’ than quantitative methods however we have shown how valuable such methods can be in both understanding patients’ lived experiences and constraining further hypotheses for quantitative investigation. By collation and interpretation of themes derived from social media, we have articulated the concerns and experiences of patients with qualitative olfactory disorders and gained a better understanding of qualitative olfactory dysfunction, learning over time how the disease progresses and how it affects eating behaviour, social interactions, and mental health.

Forums such as Facebook provide much-needed moral support where members joining immediately feel connected, feel listened to and relieved to know that they are not alone. This itself can lift spirits, playing an important first step in coming to terms with what is, for many, a relatively long-term condition.

We identified seven themes arising from the corpus of unmoderated and unprovoked comments which highlighted the emotional impact of these disorders, the fluctuating nature of olfactory dysfunction, and the heretofore undescribed symptom of ‘smell lock’ or olfactory perseveration. Common trigger foods and their olfactory characteristics were identified, the most common being coffee, meat, onions and mint/toothpaste. We noted particularly alteration of faecal odours as acquiring a more positive hedonic valence. The commonality of trigger foods and their shared characteristics promises a rich field of further investigation as to the underlying mechanisms in these sensory disorders. These themes provide the frameworks for ongoing research and therapeutic intervention in smell disorders, shedding further light on the impact of these problems but also the directions to which further research should be guided.

## Data Availability

All data for which we have permission to use is included in the manuscript.

## ACKNOWLEDGEMENTS

The authors would like to acknowledge the contribution of Aidan Kirkwood for his assistance with the social media.

## AUTHORSHIP CONTRIBUTION

Datamining, manuscript first draft and review (JP); conception, Facebook management, datamining, figures and review of manuscript (CK); manuscript writing and review (BS); manuscript writing and review (CH); conception, manuscript writing and review (SG).

